# Identifying community nurses’ contributions to end-of-life care: an online survey study

**DOI:** 10.64898/2026.05.08.26352717

**Authors:** Ben Bowers, Melissa Fielding, Emily Ashwell Massey, Efthalia Massou, Nicola Zolnhofer, Zoe Jayne, Maria Betts, Emma Clifford, Theresa Bradley, Claire McDonell, Crystal Oldman, Stephanie Lawrence, Alison Leary, Andrew Carson-Stevens, Stephen Barclay, Joodi Mourhli

## Abstract

**Background:** Demand for community-based end-of-life care is rising globally, driven by ageing populations with increasingly complex needs. Community nurses have a central role in providing end-of-life care, yet the proportion of their time spent in supporting people in their final year of life remains unclear.

**Aims:** To investigate how much of community nurses’ daily work involves caring for people in their last year of life, and the extent to which end-of-life care visits are cancelled, deferred or undertaken to unsatisfactory standards.

**Design:** Anonymous online survey and multimethod analysis.

**Setting/participants:** United Kingdom survey of community nurses, circulated via professional networks, social media and snowball sampling, between 28 April and 27 June 2025.

**Results:** A total of 1,471 nurses responded. Most worked in community and district nursing services (78.6%, 1156/1471) or specialist palliative care services (11.8%, 174/1471). Community and district nurses spent 23.5% (median) of their last shift providing end-of-life care. Over one in ten respondents (11.6% (171/1471) reported deferring at least one end-of-life visit during their last shift. Specialist palliative nurses were twice as likely to defer visits compared to community and district nurses (OR=2.48, 95% CI: 1.63–3.72, *p*<0.001). Staff shortages, demand exceeding capacity, and other systematic barriers contributed to deferring visits.

**Conclusions:** Community nurses play a vital role in end-of-life care, yet some of this important patient care is left undone or deferred. Investment in core and specialist nursing services, with efforts to enable and sustain this workforce, is urgently needed to meet globally growing demand for community-based end-of-life care.

**What is already known about the topic?:**

- Demand for community end-of-life care is growing in many countries.
- Community nurses play a key role in end-of-life care, yet the volume and complexity of their daily work supporting people in the last year of life remains poorly understood.

**What this paper adds:**

- Nurses working in community and district nursing services spent a median of 23.5% of their last clinical shift providing end-of-life care.
- Half (52%) of respondents who provided end-of-life care during their last shift reported delivering one or more aspects of this care below their professional satisfaction, due to workload and capacity issues.
- Over one in ten (11.6%) of nurses reported having deferred or cancelled end-of-life care visits on their last shift, significantly more specialist palliative care nurses (24%) than community and district nurses (10.4%).

**Implications for practice, theory or policy:**

- Our findings reveal a notable proportion of deferred and cancelled end-of-life care visits and care not undertaken to nurses’ professional satisfaction.
- Sustained, intentional investment in core and specialist nursing services, together with improved system-wide integration, is needed to support this vital workforce.
- Further research is necessary to understand how community and district nurses and specialist palliative care nurses can most effectively prioritise end-of-life care within finite resources and competing demands.

## Introduction

Healthcare systems must make optimal use of their limited community nursing workforce to meet rising demand and deliver safe, effective end-of-life care. More people are dying at older ages with increasingly complex long-term conditions and end-of-life care needs.^1–5^ The vast majority of people’s last year of life is spent at home,^6,7^ with half of deaths globally occurring at home.^8^ In 2024, 49.4% of the 568,613 deaths in England and Wales occurred in community settings.^9^ International policy and practice is increasingly shifting from hospital to community-based end-of-life care, alongside growing societal need for this essential area of clinical care.^1,2,7,8,10^

Community nurses have central roles and responsibilities in coordinating and delivering person-centred healthcare at home,^11–17^ yet their contributions to end-of-life care (care in the last year of life)^18^ remain under-recognised and under-researched.^16,17,19,20^ Community nurses working in specialist palliative care services focus predominantly on assessing and managing complex end-of-life care needs.^12,21^ Many patients die in the community supported by community and district nursing services, general practice teams, and care home staff, without requiring or receiving additional input from specialist palliative care services.^7,13,16,19^ For most nurses working in community and district nursing services and general practice, end-of-life care is only one component of their overall work; however, this clinically important patient care is prioritised by community nurses despite limited resources ^12,16,22,23^ Ensuring patients experience a period of end-of-life care and a death where symptoms are well controlled and patients are surrounded by those important to them in a familiar environment is seen as a core responsibility of community nursing and component of good care.^15,24–28^

To date, few studies have quantified the proportion of end-of-life care work undertaken by community nurses across specialities and services.^16,17,19,20,22^ In 2021-2022, 23 community and district nursing services in the United Kingdom (UK) reported spending 5.6% of clinical time providing palliative care, based on patients recorded conditions.^29^ Other data suggests between 10% and 14% of UK community and district nursing service interactions relate to providing palliative care or care to people in the last three months of life.^30–32^ A New Zealand study found 21% of patients on community and district nursing service caseloads had palliative care needs.^19^ These figures vary considerably due to different criteria used to define and measure end-of-life care work.

It also remains unclear how much end-of-life care work is deferred or is left undone as rising demand and complexity outpace limited resources.^16,17,33^ This demand verses capacity gap is reported to have widened in recent years,^19,22,34^ accelerated by changes in practice, understaffing, and persistent recruitment and retention issues.^11,16,19,22,35,36^ Community and district nurses have assumed expanded roles, including prescribing, verifying deaths and leading complex end-of-life care decisions.^16,35^ These developments are partly a result of wider changes to services. In the UK, general practitioners (family doctors) have increasingly adopted a medical consultant model of care, offering increasingly remote support and relying on nurses to visit patients, lead care planning and make clinical decisions with families.^11,16,37^ Under significant time and resource pressures, community nursing teams face ethically difficult trade-offs, at times needing to prioritise safety-critical technical nursing activities and deferring or cancelling end-of-life care visits.^12,16,22,38^

To develop sustainable system-wide workforce plans that meet growing demand, it is essential to understand the extent of end-of-life care work community nurses provide and whether aspects of this work are routinely left undone. Our study aimed to investigate how much of community nurses’ daily clinical work involves caring for people in their last year of life, and the extent to which end-of-life care visit are cancelled, deferred, or undertaken to unsatisfactory standards.

## Methods

### Study design

An online survey of community nurses working in the UK. Our data analysis employed a multimethod approach.^39^ Quantitative findings provided insights into nurses’ working practices and informed the qualitative thematic analysis, which explored nurses’ stated reasons for deferring end-of-life care. The research was underpinned by a critical realist approach.^40^

### Ethical approvals

The *[University anonymised for peer review]* Department of Engineering Ethics Committee [693/2025] approved the study. Participants provided online consent before proceeding to the survey; no personal identifiable data were collected.

### Population and setting

UK-based registered nurses, nursing associates, healthcare assistants and support workers working in any community settings were eligible to take part.

### Definitions

End-of-life care was defined as providing direct clinical input for patients considered ‘likely to be in their last year of life’^18^ during respondents’ most recent clinical shift. Community nursing service types and salary bands are defined in Table 1.

**Table 1.**
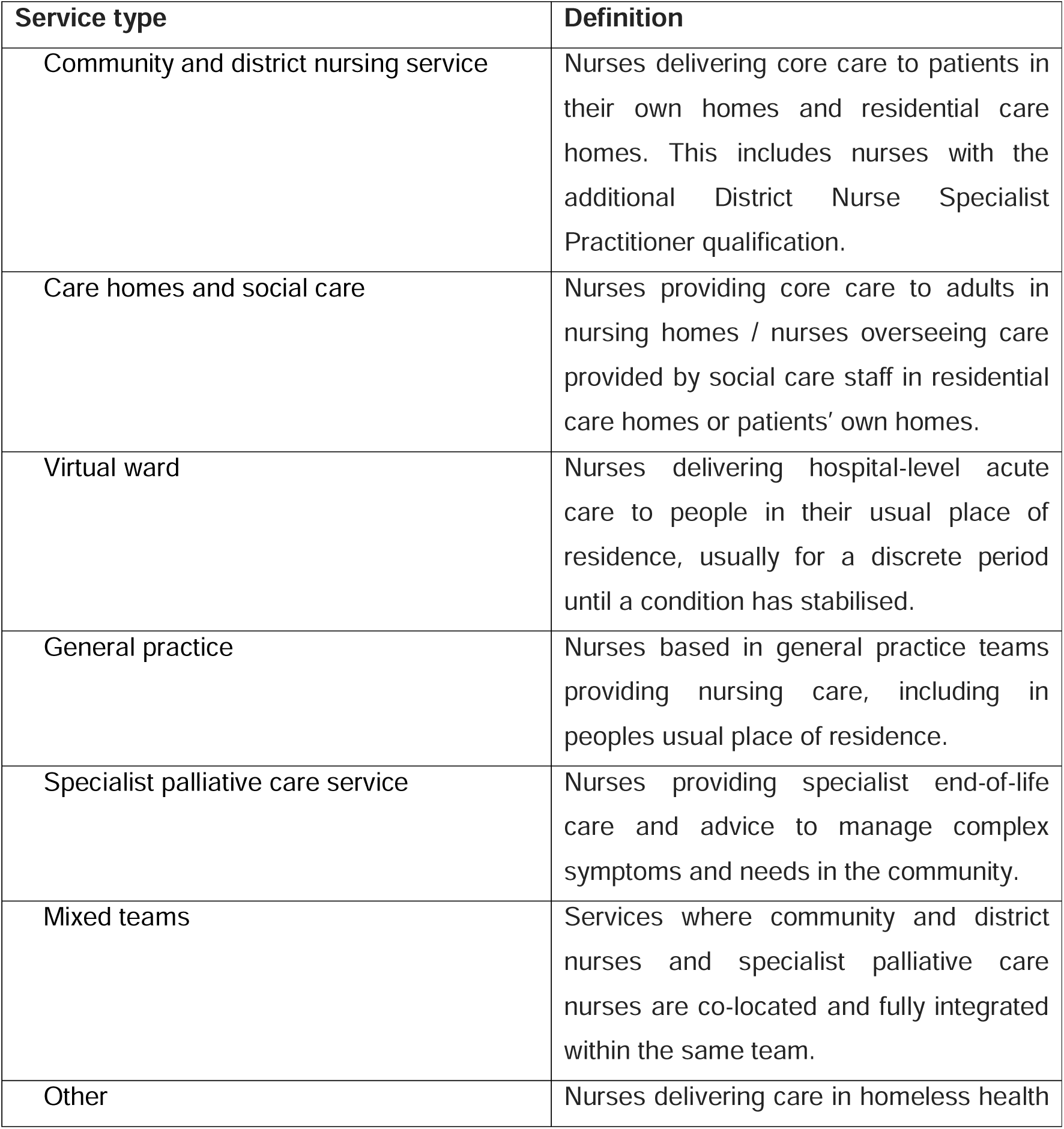
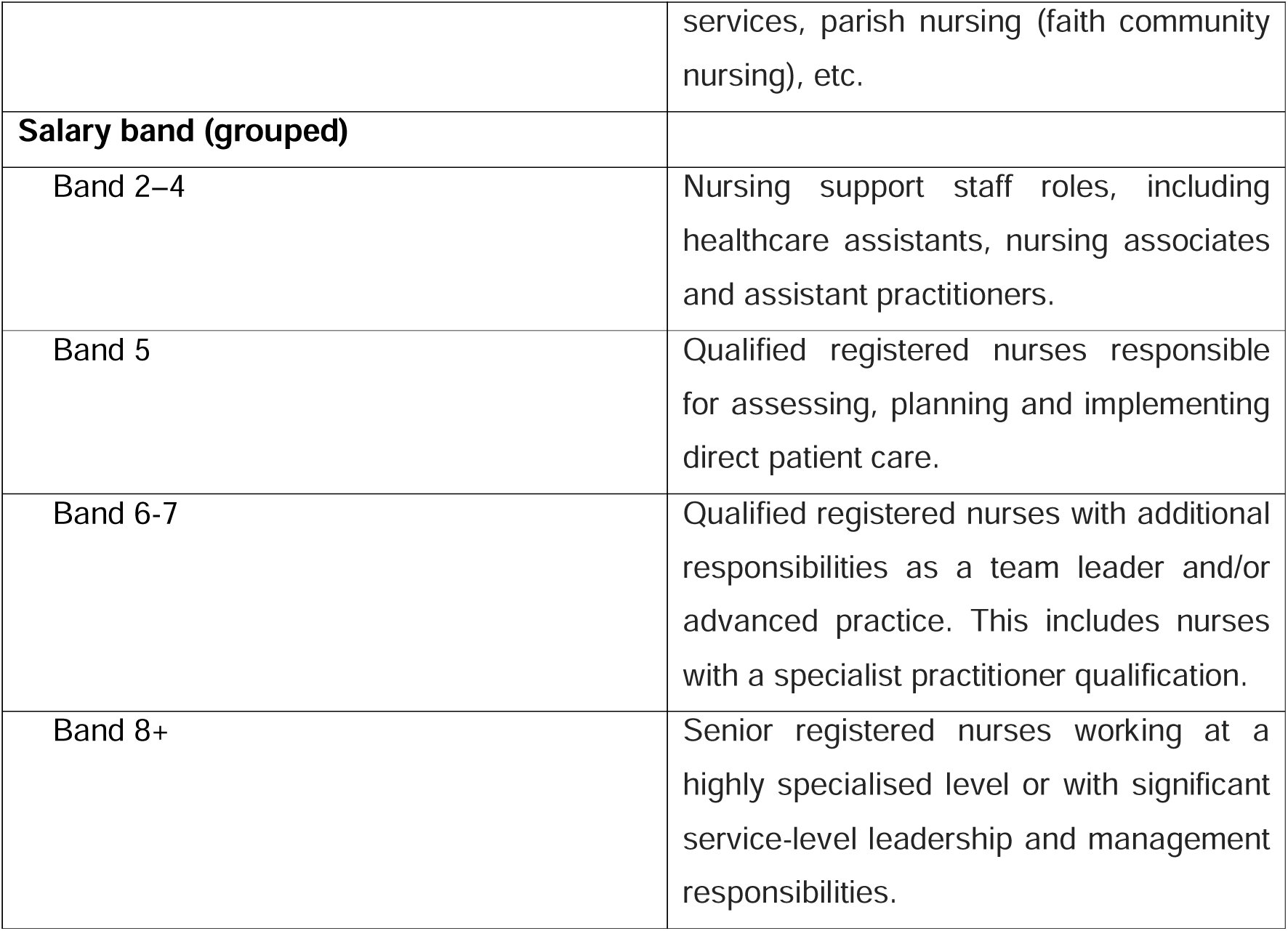
Community nursing service types and salary bands in the United Kingdom.

### Sampling and recruitment

Study information along with the survey link were disseminated via social media (LinkedIn, Bluesky / Instagram / X / Facebook), two national nursing conferences, newsletters and emails through professional networks and the research team’s existing contacts. National networks included the Queen’s Institute of Community Nursing (QICN), Royal College of Nursing District and Community Nursing Forum and Pain and Palliative Care Forum, Queen’s Nursing Institute Scotland, Association of District Nurse and Community Nurse Educators, National District Nurses Network and Marie Curie. Recipients were encouraged to forward the study material to other community nurses. This multi-pronged approach facilitated snowball sampling.

### Data collection

The online survey was hosted on Qualtrics and included quantitative items on participant demographic information, the length of their most recent shift (including time spent providing end-of-life care), and any end-of-life care that was delivered to an unsatisfactory standard, deferred or cancelled during that shift. An optional open-ended question invited participants to describe the reasons for deferring or cancelling end-of-life care visits (Supplemental Document 1). Data were collected between 28 April and 27 June 2025.

### Data analysis

Data were analysed using a multimethod approach. The quantitative findings guided the qualitative lines of enquiry, generating complementary contextual insights.^41^

Quantitative data were analysed in R (version 4.5.1) using descriptive and inferential statistical methods. Categorical data are reported as frequencies and percentages, and continuous data as medians (interquartile ranges: IQR) and means (standard deviations: SD). A minimum sample size of 900 participants was calculated *a priori* to enable statistical analysis, including Kruskal-Wallis followed by Bonferroni-adjusted Dunn’s tests, Chi-square and Fisher’s exact tests, followed by linear and logistic regression models (Supplemental Document 2). Statistical significance was set at p<0.05.

A reflexive thematic analysis approach was used to inductively analyse short free-text responses regarding reasons for the deferral or cancelation of end-of-life care visits,^42,43^ with particular attention on characteristics identified as statistically significant in the quantitative analysis. This thematic analysis was undertaken collaboratively using Microsoft Excel. MF, a critical geographer, collaborated with ZJ and NZ, experienced community nurses, to familiarise themselves with the qualitative data and generate initial codes. Through iterative group discussions and reflexive engagement with the dataset, the team refined coding and identified patterns of shared meaning in the terms commonly used by respondents. These stages were used to develop overarching themes that reflected the recurring stated reasons for deferred or cancelled visits.

## Results

A total of 1471 UK-based nurses took part in the study. Most worked in community and district nursing services (78.6%, 1156/1471) or specialist palliative care services (11.8%, 174/1471). Respondents were based across England (73%, 1074/1471), Scotland (12.2%, 180/1471), Wales (8%, 117/1471) and Northern Ireland (6.8%, 100/1471). Nearly half of respondents (48.7%, 781/1471) reported that they had worked overtime during their last clinical shift, with a median of 1 hour (IQR 0.5–1.5; range 0.5–10). On average, nurses cared for eight patients during their last clinical shift. Only 7.5% (111/1471) reported that their last clinical shift occurred during an evening or night (out-of-hours) period. See Table 2.

**Table 2.**
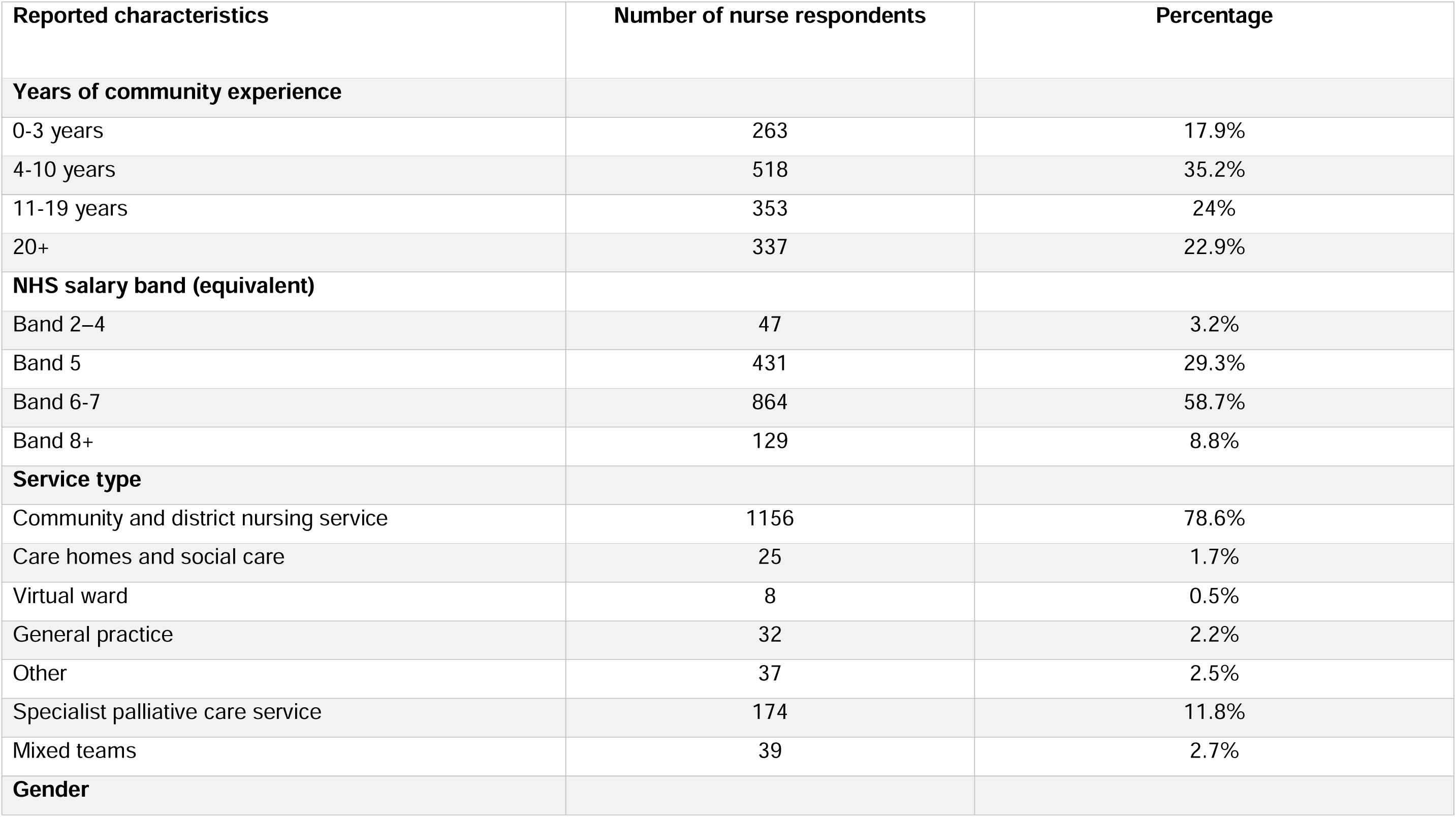
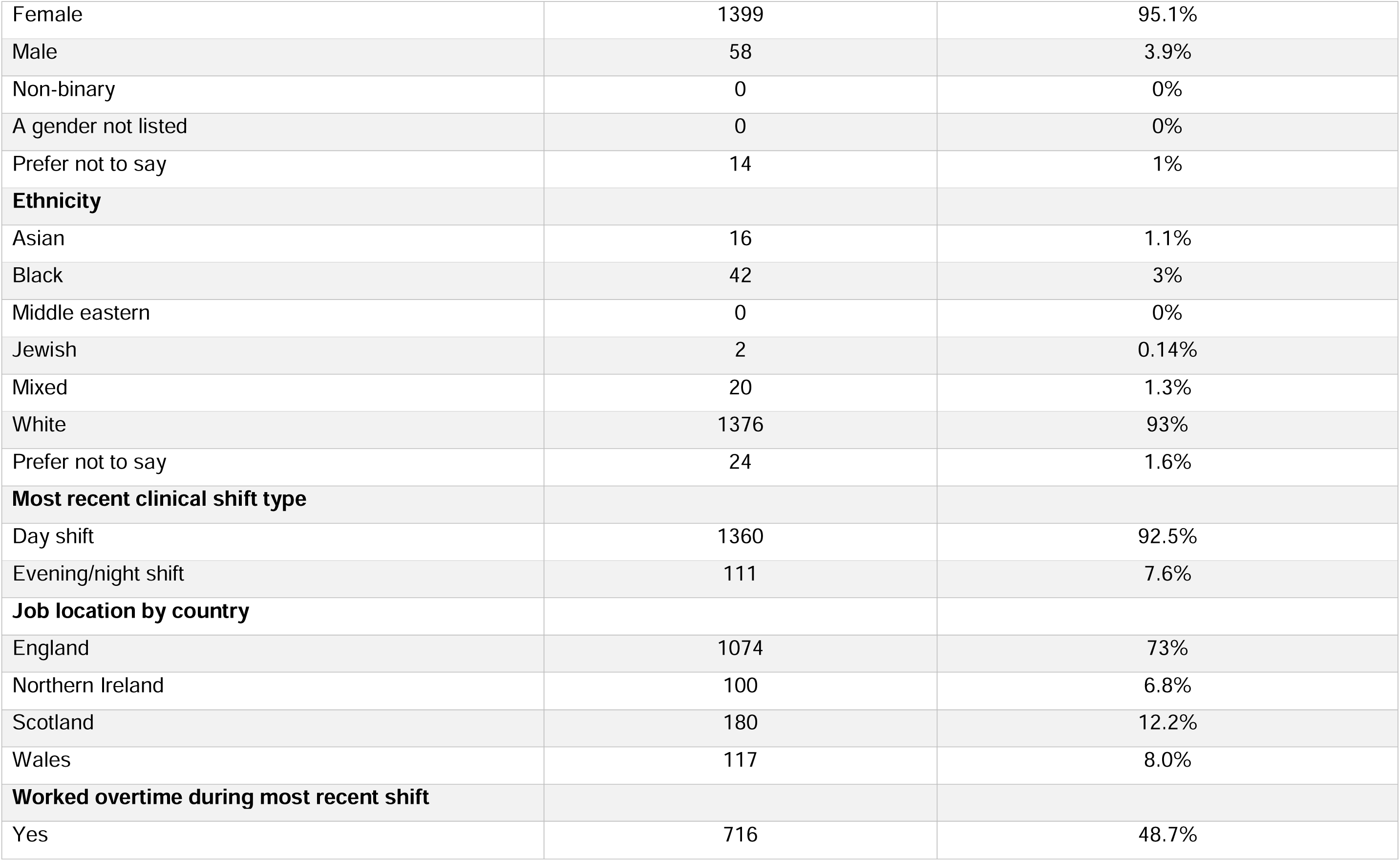
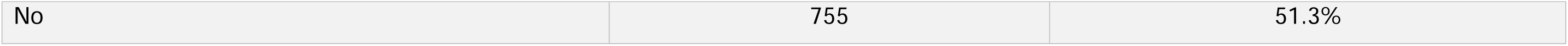
Participant characteristics and most recent clinical shift details (*n* = 1471).

| **Reported characteristics** | **Number of nurse respondents** | **Percentage** |
| --- | --- | --- |
| **Years of community experience** |  |  |
| 0-3 years | 263 | 17.9% |
| 4-10 years | 518 | 35.2% |
| 11-19 years | 353 | 24% |
| 20+ | 337 | 22.9% |
| **NHS salary band (equivalent)** |  |  |
| Band 2–4 | 47 | 3.2% |
| Band 5 | 431 | 29.3% |
| Band 6-7 | 864 | 58.7% |
| Band 8+ | 129 | 8.8% |
| **Service type** |  |  |
| Community and district nursing service | 1156 | 78.6% |
| Care homes and social care | 25 | 1.7% |
| Virtual ward | 8 | 0.5% |
| General practice | 32 | 2.2% |
| Other | 37 | 2.5% |
| Specialist palliative care service | 174 | 11.8% |
| Mixed teams | 39 | 2.7% |
| **Gender** |  |  |
| Female | 1399 | 95.1% |
| Male | 58 | 3.9% |
| Non-binary | 0 | 0% |
| A gender not listed | 0 | 0% |
| Prefer not to say | 14 | 1% |
| **Ethnicity** |  |  |
| Asian | 16 | 1.1% |
| Black | 42 | 3% |
| Middle eastern | 0 | 0% |
| Jewish | 2 | 0.14% |
| Mixed | 20 | 1.3% |
| White | 1376 | 93% |
| Prefer not to say | 24 | 1.6% |
| **Most recent clinical shift type** |  |  |
| Day shift | 1360 | 92.5% |
| Evening/night shift | 111 | 7.6% |
| **Job location by country** |  |  |
| England | 1074 | 73% |
| Northern Ireland | 100 | 6.8% |
| Scotland | 180 | 12.2% |
| Wales | 117 | 8.0% |
| **Worked overtime during most recent shift** |  |  |
| Yes | 716 | 48.7% |
| No | 755 | 51.3% |

### End-of-life care provision

The median proportion of respondents’ last shift spent providing end-of-life care was 26.3% (IQR 13.3–46.7%). Nurses working in community and district nursing services spent a median proportion of 23.5% of their last shift providing end-of-life care (IQR 12.5–40.0%). In contrast, specialist palliative care nurses spent significantly greater proportion of their shift providing end-of-life care (median 66.7%, IQR 50–87.5%) (*p* < 0.001). They also cared for more patients receiving end-of-life care per shift (median 4, IQR 3–6) than community and district nurses (median 2, IQR 1–4). See Table 3.

**Table 3:**
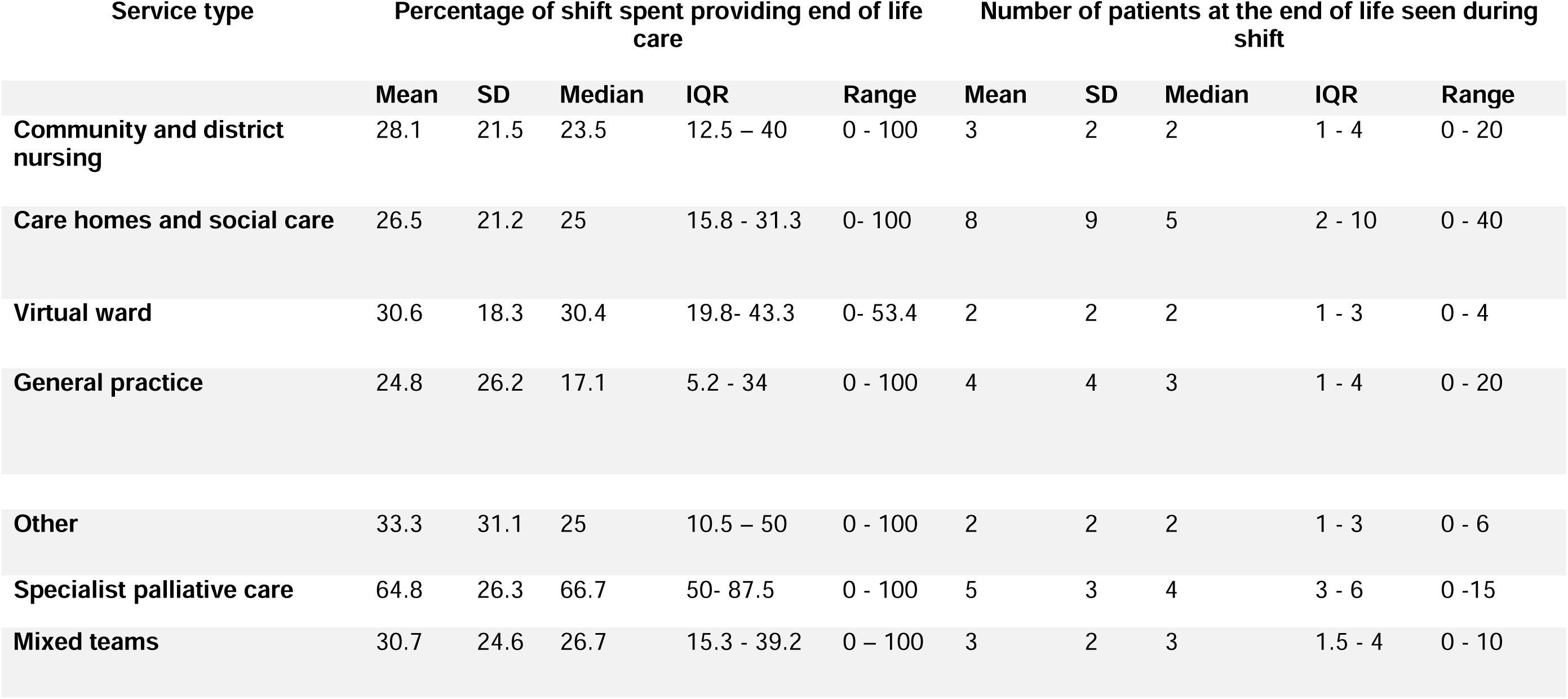
Participants’ reported end-of-life work during their most recent clinical shift by provider service type (*n* = 1471).

Across provider service types, Band 6 and 7 nurses spent significantly greater proportions of their shifts delivering end-of-life care compared with Band 5 nurses (*p* < 0.001). Nurses with 4–10 years’ experience spent significantly more time providing end-of-life care than those with 0–3 years (*p* < 0.001).

In an adjusted linear regression model, specialist palliative care nurses spent 34.8% more of their shift delivering end-of-life care than community and district nurses (95% CI: 31.1, 38.5; *p* < 0.001). Band 5 nurses across services spent 5.0% less of their shift on end-of-life care than Band 6–7 nurses (95% CI: -8, -2; *p* < 0.001), and nurses with 0–3 years of experience spent 4.9% less time on end-of-life care than those with 4–10 years (95% CI: -8.5, -1.5; *p* = 0.006) No significant difference in delivering end-of-life care were noted across other service types, pay bands, or experience levels (Supplemental Document 3).

### Injectable medication symptom control

Nearly half (45.8%, 673/1471) of respondents reported administering injectable end-of-life symptom control medications during their most recent shift. Most of these respondents worked in community and district nursing services (85%, 573/673), with a smaller proportion working in specialist palliative care services (9.9%, 67/673). Community and district nurses administered injectable medications to between 0-9 patients in their last shift (median 0, IQR 0-1); specialist palliative care nurses administered to between 0-10 patients (median 0, IQR 0-1).

The frequency of injectable medication administration differed significantly by service type, pay band, years of experience, and recent shift. Nurses working evening/night shifts administered injectable medication to more patients (median 2, IQR 1-3) than those on daytime shifts (median 0, IQR 1-2) (*p* < 0.001). Nurses with 4–10 years of community experience also reported administering injectable medications to more patients than both less or more experienced colleagues (*p* < 0.01).

### End-of-life work left undone

Over one in ten respondents (11.6%, 171/1471) reported deferring or cancelling end-of-life care visits during their last shift. Such deferrals and cancellations were significantly more frequent among specialist palliative care nurses (42/174; 24%) than among community and district nursing staff (120/1156; 10.4%) (*p* < 0.001). Band 6 and 7 nurses most frequently reported deferring or cancelling visits (*p* < 0.001), while no significant association was observed with years of experience.

In the adjusted logistic regression model, specialist palliative care nurses were significantly more likely to defer or cancel visits when compared to community and district nurses (OR=2.48, 95% CI 1.63–3.72, *p* < 0.001). Band 5 nurses were significantly less likely to cancel visits than Band 6 and 7 nurses (OR=0.49, 95% CI 0.31–0.73, p<0.001), while Bands 2-4 and Band 8 and above nurses showed no statistically significant differences.

Half (52%, 694/1334) of respondents who provided end-of-life care during their last shift identified delivering one or more aspects of this care below their professional satisfaction (Table 4). This most frequently included psychological care/support (23.2%, 309/1334) and coordination of care (14.7%, 196/1334).

**Table 4.**
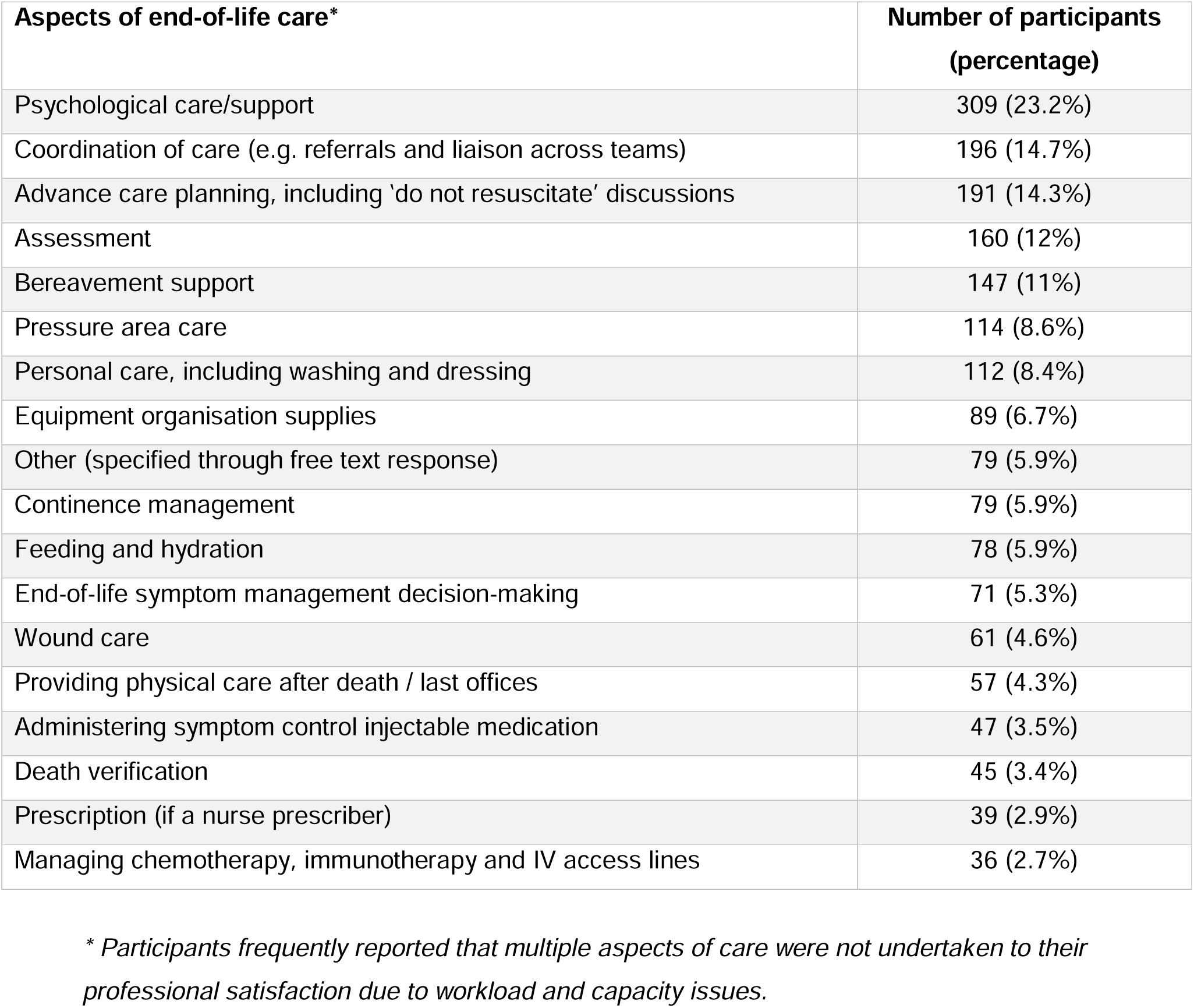
End-of-life care work not done to nurses’ professional satisfaction (*n* = 1334).

| **Aspects of end-of-life care\*** | **Number of participants**<br>**(percentage)** |
| --- | --- |
| Psychological care/support | 309 (23.2%) |
| Coordination of care (e.g. referrals and liaison across teams) | 196 (14.7%) |
| Advance care planning, including 'do not resuscitate' discussions | 191 (14.3%) |
| Assessment | 160 (12%) |
| Bereavement support | 147 (11%) |
| Pressure area care | 114 (8.6%) |
| Personal care, including washing and dressing | 112 (8.4%) |
| Equipment organisation supplies | 89 (6.7%) |
| Other (specified through free text response) | 79 (5.9%) |
| Continence management | 79 (5.9%) |
| Feeding and hydration | 78 (5.9%) |
| End-of-life symptom management decision-making | 71 (5.3%) |
| Wound care | 61 (4.6%) |
| Providing physical care after death / last offices | 57 (4.3%) |
| Administering symptom control injectable medication | 47 (3.5%) |
| Death verification | 45 (3.4%) |
| Prescription (if a nurse prescriber) | 39 (2.9%) |
| Managing chemotherapy, immunotherapy and IV access lines | 36 (2.7%) |

*\* Participants frequently reported that multiple aspects of care were not undertaken to their professional satisfaction due to workload and capacity issues.*

### Reasons for deferring and cancelling visits

Of the 171 nurses who reported deferring or cancelling end-of-life care visit during their last shift, 163 (95.3%) provided reasons for this. Most respondents worked in community and district nursing services (86.5%, 141/163) or specialist palliative care services (8.6%, 14/163). The majority of responses (89%, 145/163) related to deferring working during day shifts.

#### Demand exceeding capacity

The most frequently reported reason for deferral was insufficient workforce capacity (68.7%, 112/163), with respondents repeatedly citing high workload demands, chronic understaffing, long travel distances, limited availability of skilled personnel, and deficits in staff expertise in end-of-life symptom management. Care was reported to have become reactive rather than proactive due to the demand verses capacity gap:

> *‘[We do not have] enough staff to cope with the volume of patients requiring visits, only able to provide reactive visits for palliative patients at present.’*
>
> *District nursing sister, Band 6, England*

#### Clinical prioritisation

Respondents described scenarios where urgent or unpredictable clinical needs, including symptom crises or complex wound care, took precedence when demand outpaced capacity. This led to the postponement of less urgent end-of-life care visits. Additionally, ‘complex caseloads’ often demanded more time and coordination than could be accommodated during shifts and contributed to the need to make difficult prioritisation decisions. These trade-offs when prioritising work caused respondents ethical and emotional difficulty. Nurses repeatedly reported deferring essential but non-urgent end-of-life support:

> *‘Lamentably, we are in a position within our district nursing team of having to choose between priority / essential calls. Those patients on syringe drivers naturally are always priority, but those who are on their palliative journey and may be receiving once or twice weekly support visits are often moved.’*
>
> *District nursing sister, Band 7, Wales*

#### Logistical and systemic barriers

Logistical and systemic barriers in community settings impeded timely end-of-life care, even when workforce capacity was less pressing. Delays in access to essential medications and equipment, inadequate referrals, and poor discharge planning all contributed to delayed and deferred visits. Access to injectable medications was a recurrent concern, especially out-of-hours, with compounded impact of delayed prescribing and the challenges of locating open pharmacies with appropriate stock during evenings and weekends:

> *‘Deferred due to lack of appropriate prescribing of medications and no medications in the home. Poor planning, poor referral, delayed care. Meaning we have to wait for an out of hours GP to assess and prescribe, then families have a nightmare finding a pharmacy that… stock end-of-life medications - especially in the evening and overnight - leaving the patient waiting and suffering’*.
>
> *Community staff nurse, Band 5, England*

Other barriers included technical issues with electronic records systems and inadequate informational continuity of care across services. Few deferrals were linked to patient being unexpectedly admitted to hospital or not being at home when visited.

## Discussion

### Main findings

Our study quantifies community and district nurses’ significant daily contributions in providing end-of-life care. End-of-life care represented a median of 23.5% of respondents’ most recent shift, underscoring their role as core providers. This is notably higher than previous estimates of palliative and end-of-life care accounting for between 10% to 14% of UK-based community and district nurses’ work.^30–32^ Our findings correspond with community nurses’ accounts of their crucial contributions ^15,16,24,44^ and challenge narratives that position end-of-life care as a minor component of non-specialist practice.

Respondents frequently reported providing one or more aspects of end-of-life care below their professional satisfaction. Crucially, 11.6% of nurses deferred or cancelled at least one end-of-life visit during their last shift. Deferrals across services were driven by chronic staff shortages, demand consistently exceeding capacity, and difficult prioritisation decisions. Additional cross-organisational barriers further compounded these challenges.

### What this study adds

Taken together with wider research, our findings demonstrate that sufficient community nursing workforce capacity is critical for delivering timely, equitable and high quality community end-of-life care.^12,19,22,45,46^ Rising demand for home-based nursing care as populations age has placed significant pressure on the workforce in numerous countries including the UK and New Zealand.^12,19,22,36,47^ Pressures to reduce staffing costs in many countries providing publicly funded healthcare, combined with persistent recruitment and retention issues, have negatively impacted community nursing capacity.^11,16,22,34,48^

Recent economic analysis reinforces the case for sustained investment in a community nursing workforce that can provide core end-of-life care alongside a range of other nursing interventions, flexing to population needs. The Nuffield Trust found the average cost of a community nurse face-to-face visit is just £57 in England, compared to £287 for an emergency ambulance paramedic home visit, or £2,254 for an emergency short stay admission.^34^ These differences in costs support the economic case for investing in community nursing capacity to meet the needs and care preferences of ageing populations, particularly when proactive and responsive community end-of-life care can help to alleviate avoidable suffering and prevent unplanned hospital and hospice admissions.^7,23,34,45,49,50^

Our findings reveal a concerning proportion of deferred and cancelled visits, alongside care delivered below nurses’ professional standards. End-of-life care visits and activities that go undone in the community are easily overlooked, as staff activity planning and reporting systems do not capture missed work. As a result, it is difficult to quantify when clinically important and safety-critical care is deferred or is left undone unless patient safety incident reports are submitted. However, adverse incidents and near misses are systematically under-reported in healthcare.^51,52^ Studies consistently show that a cultural emphasis on technical task-focused care, combined with high workloads, limits community and district nurses’ abilities to provide person-centred care.^16,19,22,44,53^ When sub-standard practice and deferred care become normalised, professionals may become desensitised to the associated risks and less likely to report adverse events, including harms arising from delays in delivering end-of-life care and symptom management.^46,54,55^ Further research is necessary to understand the impact of end-of-life care left undone and how community nurses can best prioritise clinically important and safety-critical work within finite resources and competing demands.^16,22^

Community healthcare systems must identify effective strategies to integrate services, ensuring cost-effective, integrated care that optimises available workforce resources.^45^ Notably, our findings indicate that specialist palliative care nurses were nearly twice as likely to defer visits compared to community and district nurses. This difference may reflect the prioritisation of end-of-life care visits within community and district nursing teams as they often involve clinically managing last-days-of-life symptoms and providing hands-on nursing care.^12,13,19,22,23,28^ In contrast, specialist palliative care nurses frequently provide care coordination and symptom management input in the months before death and typically have more time allocated per visit,^12,24,56,57^ giving them greater flexibility to defer appointments without immediate detriment to patient care. They may also redirect patients to other healthcare professionals, including community and district nursing teams and general practitioners (family doctors), when balancing competing demands. Our findings and comparable studies highlight both service adaptability and the need for clearly defined roles, responsibilities and agreements on how to best use finite resources when core and specialist services are operating under significant pressure.^11,16,17,34,58^ In practice, there are significant opportunities for local healthcare systems to regularly review and plan the coordinated, integrated use of finite resources across services. Internationally, further research is needed to understand how core and specialist community nursing services and general practitioners can best work together to meet fluctuating workload demands without compromising person-centred end-of-life care.^11,16,19,37,45^

### Strengths and limitations

Defining end-of-life care within the survey as the provision of direct clinical input for patients considered ‘likely to be in their last year of life’ reduced ambiguity associated with the terms “palliative”, “end-of-life”, and “terminal” care.^59^ The large number of responses and participants’ diverse geographical spread across all four nations provide a robust snapshot of reported end-of-life care work across the UK. The multi-pronged recruitment approach addressed the lack of any central lists of nurses working in community settings in the UK. Previous studies reported challenges recruiting from community and district nursing services, often due to nurses lacking time or confidence to engage in research activities.^60,61^ In contrast, our recruitment approach effectively engaged nurses: the survey required around eight minutes to complete.

Self-selection bias is possible. Nurses with an interest in end-of-life care may have been more likely to participate; relatively few respondents reported delivering no end-of-life care in their last shift. Only 47 participants worked in Band 2-4 nursing support staff roles, despite these roles becoming increasingly commonplace in community nursing services.^22^ Support staff in these roles may provide less end-of-life care than Band 5 and above nurses, depending on the complexity of patient needs.^16^ Nurses working in care homes and social care were underrepresented, limiting the analysis options. Further tailored research focusing on the end-of-life care contributions of these groups is warranted. Respondents were only asked for the reasons for deferring or cancelling care; we were unable to ascertain if safety-critical visits were assigned to other services instead.

### Conclusion

This UK-wide study demonstrates that community nurses are routinely providing a very significant amount of end-of-life care, yet some of this important work is being left undone or being deferred. The end-of-life work contributions of nurses in community and district nursing services are frequently underestimated in healthcare policy and system-wide workforce plans. These findings have international relevance and transferability to other publicly funded healthcare systems: the methods could be replicated in different countries to capture the contributions of nurses across specialities and services. Addressing rising international demand for community-based care will requires sustained investment in core and specialist nursing services, alongside system-wide integration, to retain, support and develop this vital workforce.

## Data Availability

The anonymous dataset used in this study may be requested by researchers through contacting BB, the lead author, and on completion of a data use agreement.

## Acknowledgements

The authors would like to thank James Brimicombe and Cassie Hoyland, Data Manager and Data Coordinator, Primary Care Unit, Department of Public Health and Primary Care, University of Cambridge, for their expertise and help in planning and running this study. Members of the QICN Community Nursing Research Forum for their valuable helping sharing the survey widely and for talking part. Professor Fliss Murtagh, University of Hull, for her insightful advice on the quantitative analysis.

## Author Contributions

BB, SB, AL, CO and ACS designed the study. MF, ZJ NZ conducted the qualitative analysis with input from BB. JM and BB carried out the quantitative analysis with input from EM, EA, MB, EC, TB and CK. EA and JM created the visuals. All authors contributed to the interpretation of the results and the writing of the manuscript. BB, SB, SL and CO provided the resources. The corresponding author attests that all listed authors meet authorship criteria and that no others meeting the criteria have been omitted. BB is the guarantor and affirms that the manuscript is an honest, accurate and transparent account of the study being reported.

