## Supplemental Document 1 for "Identifying community nurses’ contributions to end-of-life care: an online survey study"

**Community nurses end-of-life work survey questions**

1. **How long have you worked as a nurse in the community?**

Less than 1 year

1 to 3 years

4 – 6 years

7 – 10 years

11 – 14 years

15 – 19 years

20 – 24 years

25 – 29 years

30 + years

**What is your current role?** Please tick the closest to your role even if your job title isn’t here.

Healthcare Assistant or Support Worker

Assistant Practitioner

Trainee Nursing Associate

Nursing Associate

Student Nurse (on community nursing placement)

Community Staff Nurse/Senior Staff Nurse

District Nurse (DNSPQ)

District Nurse (apprentice or student)

District Nursing Sister/Charge Nurse

Sister/Charge Nurse

Community Matron

General Practice Nurse

Advanced Nurse Practitioner / Clinical Practitioner

Clinical Nurse Specialist

Learning Disability Registered Nurse

Mental Health Registered Nurse

Children's Registered Nurse

Director of Nursing/ADON

Nurse Consultant

2. **What pay band are you currently on?** Please use equivalent if you’re not on Agenda for Change.

Band 2

Band 3

Band 4

Band 5

Band 6

Band 7

Band 8a

Band 8b

Band 8c

Other e.g. VSM or greater than Band 8c

3. **What kind of organisation / team do you work in?**

Community nursing team / district nursing (any type)

Care home / nursing home

Social care (excluding care home / nursing homes)

Virtual ward

General practice

Homeless / inclusive health

Specialist palliative care / hospice at home

Prison / justice nursing

I work across a mixture of teams

None of the above

4. **Is your care setting rural, urban or mixed in nature?**

Rural

Urban

Suburban

Mixed

5. **What type of shift was your most recent clinical shift?** Please select the most appropriate

Day shift

Twilight shift (covering the hours between 6pm to 10pm)

Night shift (including from 10pm onwards)

6. **How many hours did you work on your most recent clinical shift?** This includes working through meal breaks and doing overtime. Please round up to the nearest 30 minutes

Number selector 2 – 16 hours

(note: in half an hour increment – e.g. 2 hours, 2.5 hours, 3.0 hours, 3.5 hours…)

7. **Did you work any unpaid overtime on your most recent clinical shift?**

Yes

No

8. **If yes, how much unpaid overtime did you work on your most recent clinical shift?** Please round up to the nearest 30 minutes

Number selector 0.5 hours – 10 hours

(note: in half an hour increment – e.g. 0.5 hours, 1 hour, 1.5 hours, 2 hours, 2.5 hours…)

9. **How many patients (or their families) did you visit or have direct contact with on your most recent clinical shift?**

Number selector Nil - 40

10. **How many of those direct contacts related to patients likely to be in their last year of life?** This may include people without a formally coded end-of-life diagnosis

Number selector Nil - 40

11. **How many minutes in total did you spend providing input or support for patients in their last year of life during your most recent clinical shift?** This includes time in contact with families, liaising with other professionals, organising and recording end of life care input.

Please round up to the nearest 30 minutes

Number selector nil – 16 hours

(note: in half an hour increment – e.g. Nil, 0.5 hours, 1 hour, 1.5 hours, 2 hours, 2.5 hours…)

12. **How many patients did you administer injectable end-of-life symptom control medications to on your last clinical shift?** This includes via syringe drivers.

Number selector Nil – 15

N/A – unable to administer these medications

13. **Are there any aspects of end-of-life care you / your team were unable to undertake to your professional satisfaction due to capacity and workload issues during your last clinical shift?** Please select all that applied during your shift

Assessment

Psychological care / support

Coordinating care (e.g. referrals and coordination with other teams and services)

Advance care planning, including ‘do not resuscitate’ discussions

Prescribing medication (if you are a prescriber)

End-of-life symptom management decision making

Administering symptom control injectable medication (including via syringe pumps)

Managing chemotherapy, immunotherapy and IV access lines (e.g. flushing PICC lines)

Wound care

Pressure area care

Personal care, including washing and dressing

Feeding and hydration

Organising equipment supplies

Managing continence

Verifying death (if able to do so)

Providing physical care after death / last offices

Bereavement support

Other (please specify)

14. **Did you have to delay, cancel or defer end-of-life care visits or contacts during your last clinical shift?**

Yes

No

15. **If yes, how many visits or contacts with patients and families did you cancel or defer to another day / shift on your last clinical shift?**

Number selector 1 - 20

16. **Please could you briefly describe the reasons why this work was cancelled or deferred.** Please don’t enter any names, places or other personal information that might identity either yourself or anyone else.

Free text box – 600 characters max

17. **How often does your team have planned meetings with General Practice teams to discuss the care of patients approaching their end of life?** For example, multidisciplinary meetings / Gold Standards Framework (GSF) Meetings

Daily

Weekly

Every two weeks

Monthly

Every two months

Every three months

Never

Other (please specify)

18. **Please tell us where you work?**

England – South East

England – South West

England – London

England – East

England – Midlands

England – North West

England – North East and Yorkshire

Northern Ireland (all)

Scotland (all)

Wales (all)

Channel Islands

Isle of Man

19. **What age are you?**

18 to 24

25 to 34

35 to 44

45 to 54

55 to 64

65 or older

Prefer not to say

20. **Which of the following options most closely align with your gender?**

Woman

Man

Non-binary

A gender not listed

Prefer not to say

#### 21. **How would you describe your ethnicity?**

Asian - Indian

Asian - Pakistani

Asian - Bangladeshi

Asian - Chinese

Asian - Filipina/Filipino

Asian - Any other Asian

Black - African

Black - Caribbean

Black - Any other Black background

Middle Eastern - Arab

Middle Eastern - Any other Middle Eastern background

Jewish

Mixed - White and Caribbean

Mixed - White and Black African

Mixed - White and Asian

Mixed - Any other Mixed/Multiple ethnic background

White - British

White - Irish

White - Gypsy or Irish Traveller

White - Any other White background

Prefer not to say
