## Supplemental Document 2 for "Identifying community nurses’ contributions to end-of-life care: an online survey study"

**Statistical tests conducted**

Chi-square tests of proportions were used to explore whether (i) cancelled or deferred end-of-life care visits and (ii) end-of-life care delivered below professional satisfaction were statistically associated with organisation type, pay band, and years of experience. Where assumptions for Chi-square were not met (e.g., low cell counts), Fisher’s exact test was used, and in some cases variable categories were combined or the analysis was restricted to specific categorical levels.

Kruskal-Wallis rank sum tests were conducted to investigate whether (i) the percentage of shift spent on end-of-life care and (ii) the frequency of injectable medication administration varied significantly by organisation type, pay band, and years of experience among nurse respondents. As each independent variable included multiple categories, post hoc pairwise comparisons using Bonferroni-adjusted Dunn’s tests were also performed.

Linear regression models were employed to investigate the relationship between (i) the percentage of respondents’ shifts spent providing end-of-life care; and (ii) the frequency of injectable medication administration, with predictors including organisation type, pay band, and years of experience.

Logistic regression models were used to examine the relationships between the same predictors and respondents’ reports of deferral or cancellation of end-of-life care visits (yes/no binary outcome).
